# Differences in centre of mass measurements between markerless and marker-based motion capture systems during balance and mobility assessments in individuals with chronic and sub-acute stroke

**DOI:** 10.64898/2026.02.18.26346541

**Authors:** Nigel Majoni, Elizabeth L. Inness, David Jagroop, Cynthia J. Danells, Avril Mansfield

## Abstract

Centre of mass (COM) is a key measurement used to assess balance and mobility. Marker-based motion capture systems have traditionally been used to measure COM, but they are time-consuming and prone to marker error. Markerless motion capture systems offer a potential alternative, reducing setup time while maintaining accuracy. The ease of collecting markerless data may be particularly beneficial when study participants have limited mobility, such as those with stroke. This study aimed to determine the differences in COM measurements between marker-based and markerless motion capture systems during balance and mobility tasks in individuals with sub-acute stroke. Seventeen participants completed the following tasks: walking, quiet standing, sit-to-stand, rise on toes, and backward reactive stepping. COM data were analyzed using two markerless models, a ‘default’ with 17 segments and a ‘fit’ model with 11 segments to match the marker-based model to be compared as the reference. The results showed high correlations (R^2^ = 0.75 to 0.999) and low root-mean-square differences (< 2 cm) in the anterior-posterior and medial-lateral directions. Larger differences (> 4 cm) were observed in the superior-inferior direction, particularly with the default model. These findings suggest that markerless motion capture can be used to measure COM in people with stroke, and that model selection plays an important role in COM estimates.

## 1. INTRODUCTION

Marker-based motion capture has been the gold-standard for measuring balance and gait related metrics such as centre of mass (COM).(1–3) However, applying reflective markers on individuals is time-consuming, increasing the duration of data collection, and possibly impeding natural movement patterns due to the encumbrance of equipment.(4) There is also the risk of the reflective markers falling off or moving during data collection, which impacts accuracy of COM estimates. Even small errors in COM detection can impact the interpretation of the results.(5) Recent advancements in markerless motion capture have shown to be an alternative solution in reducing the duration of data collection, while maintaining the reliability and ‘accuracy’ of conventional motion capture systems, by using deep learning algorithms to estimate joint centres directly from video data.(6,7) Specifically, markerless motion capture systems have shown comparable results to marker-based motion capture systems when assessing whole body kinematics.(6–8). To date, only one study used markerless motion capture for static balance tasks; the study observed movement strategies during these tasks, but did not evaluate other balance metrics such as COM.(9) A different study validated markerless motion capture systems compared to conventional marker-based motion capture in balance-related measurements during dynamic balance tasks.(10) That study found that COM position and motion were comparable between the markerless and marker-based systems. However, both studies were conducted in young healthy adults, which raise concerns of accuracy and usability when applied to a wider demographic (such as those with mobility deficits).

People with stroke may experience impaired balance control, demonstrate atypical movement patterns, and/or wear assistive devices (e.g. ankle-foot orthoses), which may be misinterpreted by standard pose-estimation algorithms and interfere with the markerless systems’ ability to detect the limbs. People who have previously experienced a stroke have an increased risk of falling compared to their age-matched counterparts.(11) The main contributor to the increased risk of falling post-stroke is impaired balance control.(12) Stroke-related balance impairment leads to dependence in activities of daily living, increased fear of falling and decreased quality of life.(12–14) Previous literature has found that in people who have experienced a stroke, the best predictor of return to independent living is improved balance.(15) Therefore, the validity and reliability of markerless motion capture in assessing balance related metrics in people with stroke needs to be established.

The objective of this study was to evaluate the differences between marker-based and markerless systems in measuring COM during balance and mobility assessments, and to determine whether a markerless motion capture system can be used to measure COM in people with sub-acute and chronic stroke.

## 2. MATERIALS AND METHODS

### 2.1 Study setting and apparatus

The study was conducted at KITE, Toronto Rehabilitation Institute, University Health Network in Toronto, Ontario, Canada. All data were collected in the ‘FallsLab’. Participants wore a safety harness attached to a robotic gantry overhead to allow free movement and prevent falls to the floor. We collected 3D kinematic data using a markerless motion capture system (Theia Markerless, Inc., Kingston, ON, version: 2023.1.0.3161) and a marker-based motion capture system (Vicon Nexus, Vicon, Oxford, UK).

### 2.2 Study design

The study is a validation study to assess the difference in centre of mass (COM) measurements between a marker-based (Vicon Nexus, Vicon, Oxford, UK) and a markerless (Theia Markerless, Inc., Kingston, ON, version: 2023.1.0.3161) motion capture system.

### 2.3 Participants

The sample for this validation study was drawn by convenience from participants who had already completed both marker-based and markerless motion-capture assessments in a prior study.(16) Chaumeil et al completed a similar comparison of balance measures between markerless and traditional motion capture with 16 participants.(10) We included 17 participants in our cohort which included 12 men and 5 women (Table 1). One participant wore an ankle foot orthosis, and another wore an arm sling during data collection.

**Table 1.** Participant demographics.

|  | **Male**<br>**(n=12)** | **Female**<br>**(n=5)** |
| --- | --- | --- |
| **Age (years)** | 55.4 (17.6) | 54.6 (18.9) |
| **Height (m)** | 1.76 (0.06) | 1.62 (0.06) |
| **Mass (kg)** | 85.4 (17.4) | 66.4 (14.3) |
| **Time post-stroke (days)** | 328.1 (46.2) | 171.8 (107.5) |
| **mini-BEST score** | 23.6 (2.1) | 24.6 (1.7) |
| **TUG score** | 8.27 (0.96) | 7.20 (1.10) |
| **Chronic stroke** | 12 | 2 |
| **Subacute stroke** | 0 | 3 |

### 2.4 Study protocol

Participants in the previous study completed three data collection sessions in FallsLab: pre-training, post-training, and 6-month follow-up. At the start of each session, height and body mass were measured, and participants were outfitted with 67 reflective markers; 5 markers placed on the head, 11 on each arm, 4 on the chest, 4 on the pelvis, 10 each leg, and 6 on each foot. Participants then completed the mini–Balance Evaluation System Test (mini-BEST)(17) and a gait assessment. The gait assessment included 4 perturbed walking trials (where the platform moved forward or backward suddenly on heel strike or toe off) and 6 unperturbed walking trials, presented in an unpredictable sequence. Markerless and marker-based motion capture data were collected synchronously at 100 Hz during the trials. For the purpose of this study, we analysed data from one session per participant for the 6 unperturbed walking trials and mini-BEST tasks where the markerless system is generally able to capture the full body (e.g., participant standing in the middle of the room, no occlusion from the research assistant): sit-to-stand, rise on toes, backward reactive step, and quiet standing. Demographic and stroke characteristics (e.g., time post-stroke) were obtained from chart review or directly from the participant.

### 2.5 Data processing

Three biomechanical models were created: two markerless, a ‘default’ and ‘fit’ Theia model and one marker-based model in Visual3D (C-motion, Germantown, USA, v2021.11.3). The marker-based model was the reference model for the study as marker-based motion capture has typically been used for kinematic analysis. The marker-based biomechanical model uses anthropometric data (18) and contains 11 segments: the head and trunk, left and right thighs, left and right shanks, left and right feet, left and right upper arms, and left and right forearms and hands. The ‘default’ Theia biomechanical model includes 17 segments: head, thorax, pelvis, left and right thighs, left and right shanks, left and right feet, left and right upper arms, left and right forearms, left and right hands, and left and right toes. We also created a ‘fit’ Theia model that used the same segments and anthropometry as the marker-based model. Data were filtered using a dual-pass Butterworth filter with a cut-off frequency of 6 Hz after calculating the COM in Visual 3D. COM position was calculated in 3 axes (antero-posterior, medio-lateral, and superior-inferior).

### 2.6 Data analysis

The primary measures for system validation were coefficient of determination (R^2^) for correlation and the root-mean-square difference (RMSD) across all tasks.

R^2^ was used to measure the strength of the relationship between the markerless and marker-based systems. R^2^ has been used in previous validation studies to quantify how closely the signals match in capturing human movement. RMSD provided insight into the overall differences present between the two motion capture systems.(6,10) R^2^ and RMSD were calculated in each axis.

## 3. RESULTS

A sample trial from a participant’s walking task can be seen in Figure 1, showing the signals from the three models.

**Figure 1.**
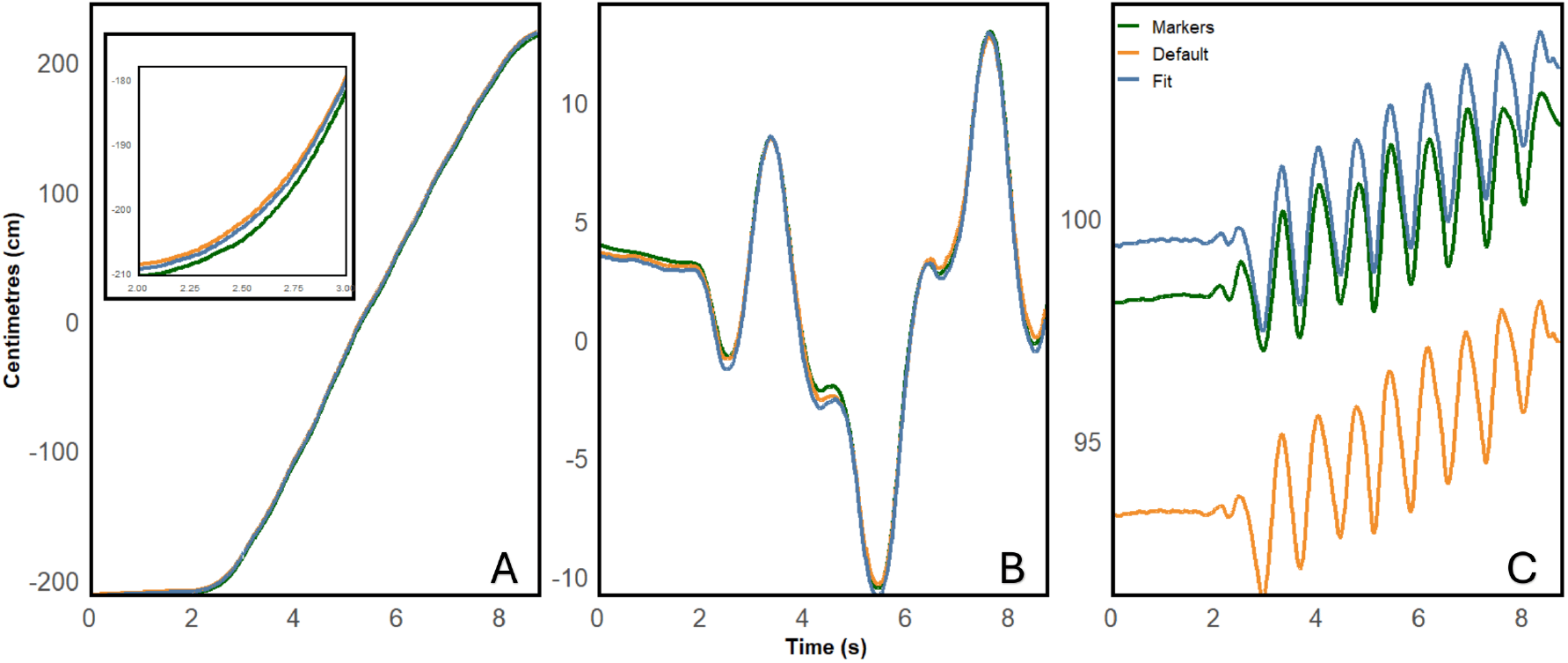
COM position in three directions A) anterior-posterior, B) medial-lateral, and C) superior-inferior, during a walking trial from a sample participant. The green line represents the reference marker-based model signal, and the orange and blue lines showcase the ‘default’ and ‘fit markerless models, respectively.

**Figure 2.**
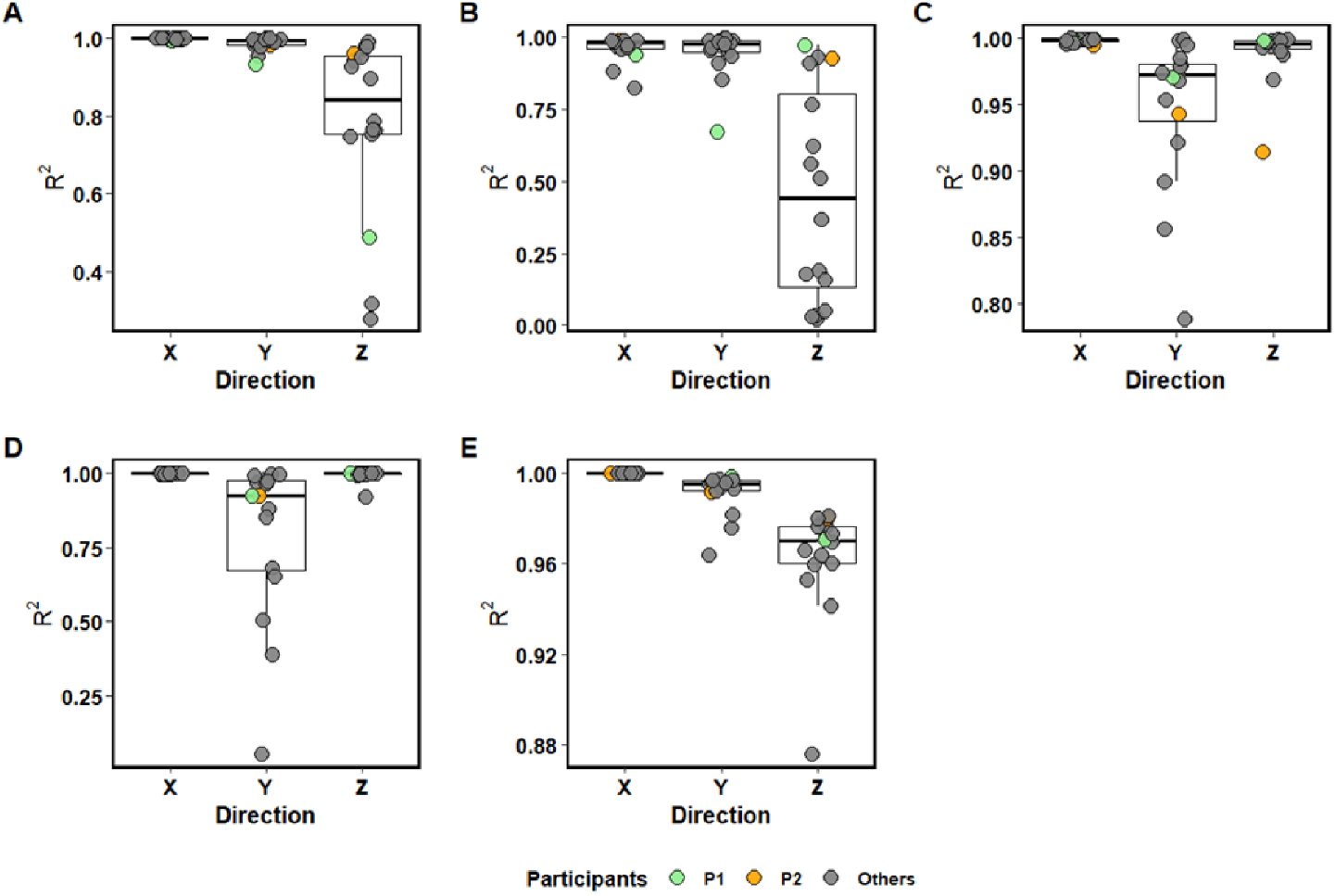
Boxplots show the R^2^ for the ‘fit’ vs. marker-based COM analysis across three directions anterior-posterior (X), medial-lateral (Y), and superior inferior (Z) for each of the five tasks: (A) backward reactive step, (B) quiet standing, (C) rise on toes, (D) sit-to-stand, and (E) walking. P1 (light green) wore an ankle-foot orthosis (AFO), while P2 (orange) wore an arm sling throughout testing; other participants are grey.

### 3.1 Comparison of the ‘default’ markerless COM model to the marker-based COM model

The RMSD in the AP direction ranged from 1.33 to 2.90 cm, and in the ML direction ranged from 0.43 to 0.58 cm. The correlations observed were high in this analysis, ranging from 0.782 to 0.999. In the SI direction, we observe a larger difference between the COM models, with RMSD ranging from 4.27 to 6.16 cm and correlations ranging from 0.422 to 0.994 across all tasks (Table 2).

**Table 2:** R^2^ and RMSD of the “Default’ vs Markers analysis for anterior-posterior (AP), medial-lateral (ML), and superior-inferior (SI) and for each task. Values presented are means with standard deviations in parentheses.

| Task | AP |  | ML |  | SI |  |
| --- | --- | --- | --- | --- | --- | --- |
| | RMSD (cm) | $R^2$ | RMSD (cm) | $R^2$ | RMSD (cm) | $R^2$ |
| Sit-to-stand | 2.90 (0.93) | 0.999 (0.001) | 0.73 (0.49) | 0.782 (0.280) | 4.27 (1.26) | 0.994 (0.020) |
| Rise on toes | 1.67 (0.93) | 0.997 (0.003) | 0.47 (0.48) | 0.951 (0.050) | 6.17 (1.04) | 0.990 (0.015) |
| Backward reactive step | 2.64 (0.85) | 0.998 (0.003) | 0.58 (0.39) | 0.983 (0.024) | 5.92 (0.94) | 0.751 (0.243) |
| Quiet standing | 1.33 (0.85) | 0.949 (0.054) | 0.43 (0.38) | 0.943 (0.094) | 5.86 (1.02) | 0.422 (0.354) |
| Walking | 2.02 (0.80) | 0.999 (0.001) | 0.49 (0.42) | 0.990 (0.015) | 5.08 (0.90) | 0.964 (0.031) |

### 3.2 Comparison of the ‘fit’ markerless COM model to the marker-based COM model

The RMSD between COM positions in the antero-posterior (AP) direction ranged from 1.01 to 1.88 cm, and in the medio-lateral (ML) direction ranged from 0.48 to 0.57 cm across all tasks. These differences were also associated with high correlations ranging from 0.783 to 0.999 in the AP and ML directions. The largest differences observed were in the superior-inferior (SI) direction, ranging from 1.10 to 1.88 cm, with high correlations ranging from 0.989 – 0.999 (Table 3). Summary results of the two participants who wore assistive devices are presented in Figures 3 and 4 to visually see how they differed from the other participants.

**Table 3.** R^2^ and RMSD of the ‘Fit’ vs Markers analysis for anterior-posterior (AP), medial-lateral (ML), and superior-inferior (SI) and for each task. Values presented are means with standard deviations in parentheses.

| Task | AP |  | ML |  | SI |  |
| --- | --- | --- | --- | --- | --- | --- |
| | RMSD (cm) | $R^2$ | RMSD (cm) | $R^2$ | RMSD (cm) | $R^2$ |
| Sit-to-stand | 1.34 (0.70) | 0.999 (0.001) | 0.73 (0.47) | 0.796 (0.267) | 1.38 (1.35) | 0.994 (0.019) |
| Rise on toes | 1.27 (0.89) | 0.998 (0.002) | 0.53 (0.46) | 0.948 (0.057) | 1.10 (0.92) | 0.989 (0.021) |
| Backward reactive step | 1.88 (0.77) | 0.999 (0.002) | 0.57 (0.37) | 0.986 (0.018) | 0.97 (0.82) | 0.783 (0.225) |
| Quiet standing | 1.01 (0.68) | 0.963 (0.045) | 0.46 (0.36) | 0.946 (0.080) | 1.03 (0.92) | 0.453 (0.357) |
| Walking | 1.47 (0.78) | 0.999 (0.001) | 0.48 (0.42) | 0.991 (0.014) | 1.20 (0.69) | 0.963 (0.029) |

**Figure 3.**
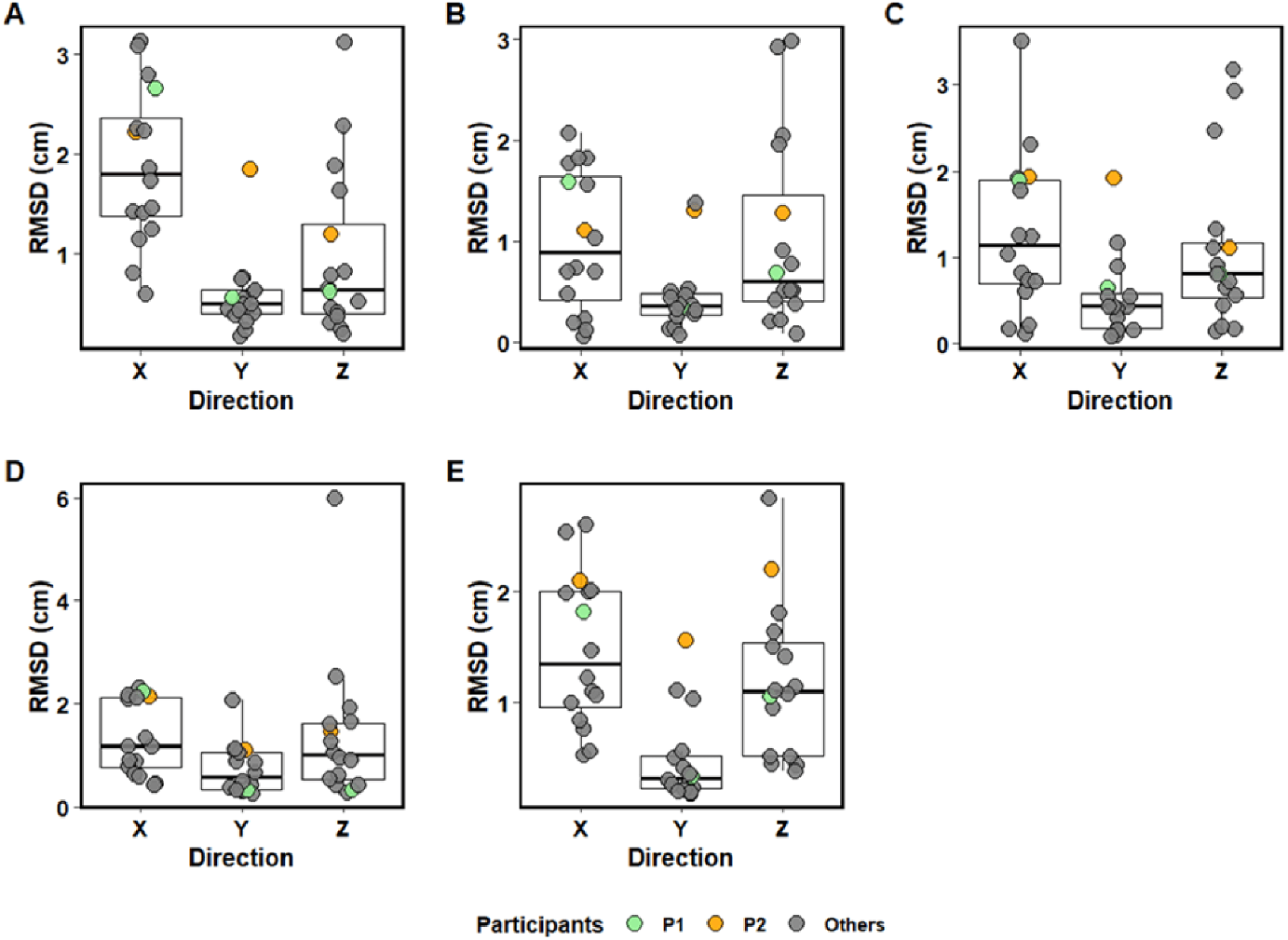
Boxplots show the RMSD (cm) for the ‘fit’ vs. marker-based COM analysis across three directions anterior-posterior (X), medial-lateral (Y), and superior inferior (Z) for each of the five tasks: (A) backward reactive step, (B) quiet standing, (C) rise on toes, (D) sit-to-stand, and (E) walking. P1 (light green) wore an ankle-foot orthosis (AFO), while P2 (orange) wore an arm sling throughout testing; other participants are grey.

## 4. DISCUSSION

This study aimed to examine differences in COM estimation between two markerless COM models and a marker-based COM model during balance and walking assessments in individuals with sub-acute stroke. Specifically, we investigated directional differences in COM position between the markerless and marker-based models. Compared to the marker-based model, both markerless models demonstrated strong correlations across all movement tasks, except during quiet standing. The most notable difference between the marker and markerless models appeared in the superior-inferior direction. These differences primarily reflect variations in the models rather than systematic bias relative to the marker-based system.

A recent study that compared COM position between markerless motion capture and marker-based motion capture, reported R^2^ to be between 0.98 – 0.99 for a walking task,(10) similar to our findings. This study also reported RMSD of 0.66, 0.28 and 0.98 in the AP, ML, and SI directions, respectively, during walking,(10) similar to our results with the ‘fit’ COM model. We observed lower correlations during quiet standing in both analyses. We attribute this finding to a high degree of noise relative to the signal in both systems. Because participants remained stationary during this trial, small deviations likely contributed to the reduced correlations between the markerless and marker models. This explanation is supported by the low RMSD in the SI direction of 1.03 cm, a difference similar to other tasks that demonstrated strong correlations in the ‘fit’ versus marker analysis. This noise-to-signal problem is inherent to quiet standing tasks: when participants remain stationary, movement signals become very small, making measurement systems highly sensitive to minor deviations and noise artifacts. This finding aligns with previous validation studies of markerless motion capture systems,(19) which similarly observed reduced correlations during quiet stance tasks.

Data were collected as part of a larger study, and the markerless system was introduced partway through the main study. To maintain consistency in the main study, we did not modify the marker set to match the markerless model. Some of the differences observed may be attributed to differences between the models and segment definitions, particularly for the analysis of the ‘default’ model vs the marker-based model. Greater agreement between the two systems could have been observed with a marker set that more closely matched the ‘default’ markerless model. We accounted for differences between models by adjusting the markerless model to more closely match the marker-based system.

For the two participants who wore the ankle foot orthosis and the arm sling we also wanted to observe whether their data differed from the other participants. We see that the participants who wore an arm sling had poorer results with greater RMSDs across most of the tasks analyzed. This difference however may not be attributed to the arm sling, rather the placement of the markers due to the interference of the arm sling. Future studies should evaluate the effects of braces and arm slings.

To participate in the main study, participants had to be able to stand for at least 30 seconds without holding onto anything and walk without a gait aid for at 10 metres at a time without stopping. Therefore, the sample of people with sub-acute and chronic stroke that was included in this study were, on average, high functioning in terms of mobility and overall impairment. Participants exhibited high balance and mobility function typical of community-ambulating individuals, with mean Mini-BEST scores of 23.6–24.6 (1.7–2.0). We also see Timed Up and Go (TUG) times of 7.2–8.3s which are comparable to healthy adults. Future studies should evaluate accuracy of markerless motion capture among people with greater motor deficits.

## 5. CONCLUSION

COM estimates from markerless motion capture have good correlations and low RMSD compared to estimates obtained from a marker-based system, suggesting that markerless motion capture can be used to assess balance and walking for people with sub-acute and chronic stroke.

## Data Availability

Participants did not consent to sharing data outside the research team.

## Acknowledgements

Funding for this project is provided by the Heart and Stroke Foundation of Canada, the Canada Brain Research Fund (CBRF), an innovative arrangement between the government of Canada (through Health Canada) and Brain Canada Foundation, the Heart and Stroke Foundation Canadian Partnership for Stroke Recovery, and the Canada Foundation for Innovation.

